# A flexible and high-throughput genotyping workflow tracked the emergence of SARS-CoV-2 variants in the UK in 2022

**DOI:** 10.1101/2023.06.03.23289684

**Authors:** Suki Lee, Stefan Grujic, Sam Modern, Angela Wann, Donald Fraser, Benita Percival

## Abstract

In late 2021, the Omicron SARS-CoV-2 variant spread rapidly worldwide. To track its emergence, and the continued evolution of SARS-CoV-2 while giving actionable epidemiological data that informs public health policy, we developed a high-throughput, automated, genotyping workflow that pairs flexible liquid handling with a re-configurable LIMS system. This workflow facilitated the real-time monitoring of the spread of BA.4 and BA.5, and by the time of its retirement, the system was responsible for typing *c*. 400,000 SARS-CoV-2 samples. When combined with a population-scale testing program, genotyping assays, can offer a rapid and cost-effective method of determining variants and horizon-scanning for changes in the pool of circulating mutations. Strategies to prepare diagnostics infrastructure for Pathogen X should consider the development of flexible systems with interchangeable components that can be rapidly re-configured to meet uncertain and changing requirements.

## 1 Introduction

The COVID-19 outbreak, caused by the severe acute respiratory syndrome coronavirus 2 (SARS-CoV-2), was declared a pandemic by the World Health Organization (WHO) in March 2020. Since then, COVID-19 waves have repeatedly overwhelmed global healthcare systems, causing over six million deaths world-wide (Dong et al., 2020).

Viral RNA dependent RNA polymerases are inherently error-prone, causing mutations to arise in the genome. Whilst SARS-CoV-2 possesses exonuclease activity that proofreads newly synthesised RNA and removes any mis-incorporated nucleotides, this process is not always accurate meaning some mutations can escape correction (Robson et al., 2020; Lauring and Hodcroft, 2021). When combined with internal selection pressures from the host’s immune system, infected individuals, this has led to the emergence of distinct SARS-CoV-2 lineages in late 2020 and early 2021 (Otto et al., 2021). Spike gene mutations, including single nucleotide polymorphisms (SNPs) P681R, E484K, K417T/N and Q493R have resulted in antigenically distinct variants. Throughout the pandemic, such variants have been named according to the backronymic nomenclature proposal Phylogenetic Assignment of named Global Outbreak Lineages (PANGOLIN) (Rambaut et al., 2020). Several variants attracted attention due to their association with increased transmissibility, reduced antibody recognition or vaccine resistance. Specifically, these include B.1.1.7 (Alpha variant), B.1.351 (Beta variant) and B.1.617.2 (Delta variant) (Krause et al., 2021). In November 2021, lineage B.1.1529 (Omicron variant) was detected in multiple countries and by March 2022, it had become the dominant strain around the world (WHO, 2022). Over 50 mutations were identified for this variant, with 32 located in the spike protein (Islam et al., 2022).

The use of whole genome sequencing (WGS) in epidemiological analysis has been demonstrated to be a robust method for reducing the impact of disease outbreak (Stevens et al., 2017). Through the COVID-19 Genomics UK Consortium (COG-UK), the United Kingdom undertook a population-wide approach to WGS surveillance of SARS-CoV-2 mutations. However, WGS is costly and entails a long data collection period due to library preparation steps coupled with bioinformatics analyses that demand high computing power (Chrystoja and Diamandis, 2014). With significant investment in infrastructure, COG-UK report that over the course of the pandemic, the sequencing capacity across all contributing sites ranged from 5k to 30k samples per week, with sequencing costs *c*. £56 - £40 sterling per sample. Average turn around times from sample receipt to sequence data upload were also reportedly highly variable, but improved from twenty days in April 2020 to six days by June 2021 (Marjanovic et al., 2022). Comparatively, PCR-based genotyping is a rapid and relatively low-cost alternative, which uses infrastructure available in a typical molecular laboratory. Such approaches could be used to pre-screen samples for sequencing and to monitor the prevalence of specific, known mutations.

Here, we report on the development and evaluation of a high throughput Reverse Transcription-quantitative Polymerase Chain Reaction (RT-qPCR) genotyping assay workflow to differentiate between multiple SARS-CoV-2 variants. The frequent emergence of new SARS-CoV-2 variants meant that valuable characteristics of any assay workflow are: high throughput, flexibility, and the rapid evaluation and deployment of new molecular reagents. These features are challenging to accommodate, even as a Research-Use-Only (RUO) assay, within a highly regulated clinical diagnostic environment. We therefore developed a robotic laboratory workflow integrated with an in-house developed laboratory information management system (LIMS). This system ensured sample and result integrity whilst providing a re-configurable and traceable workflow. Simultaneously, the platform maximised throughput and minimised spurious data input or omissions. This united platform, dubbed ReflX (RE-configurable, FLeXible), was used to track and inform real time public health decision making during the emergence of Omicron and associated sub-lineages in the United Kingdom.

## 2 Materials and Methods

### 2.1 Workflow and Integrated LIMS Solution

#### 2.1.1 Requirements

To address the challenge of screening for SNPs associated with SARS-CoV-2 variants of concern (VoC) at a population scale, a partially automated high-throughput system was developed. The requirements were:

1. A flexible integrated workflow that could screen a range of SNP targets,
2. An Information System that tracks samples and controls end-to-end, and gives full traceability of reagents, operators and equipment.
3. Utilisation of existing robotic liquid handlers and resources.
4. Workflow solution to be delivered in time for the winter COVID-19 wave of 2021-2022.
5. Flexible positive and negative controls
6. System to cherry pick and screen only SARS-CoV-2 detected samples.
7. Open-platform workflow that is reagent and labware agnostic.
8. Robust enough to be operable by personnel with minimal training.
9. Compliance with ISO15189.
10. Compliance with CFR21 Part 11.

#### 2.1.2 Laboratory Workflow Design

A summary of the laboratory workflow solution design is as follows:

1. Cherry-pick patient RNA samples called as SARS-CoV-2 Detected (using the LGC Biosearch Technologies End-Point PCR (ePCR) system) from source 96-well microtitre plate (hereafter referred to as an ELUTE plate) to destination 96-well microtitre plate (hereafter referred to as a CHERY plate) using a Hamilton MicroLab STARlet Co-RE 12-channel liquid handler. Leave two random wells empty for the negative controls, and one well in a fixed position (H12) left empty for the positive control.
2. Prepare PCR mastermix(es) according to manufacturer’s instructions.
3. Dispense, using an SPT Labtech Dragonfly Discovery liquid handler, the prepared PCR mastermix(es) into a 384-well PCR microtitre plate in 1 × 4, 2 × 2 or 4 × 1 checkerboard patterns.
4. Transfer positive controls and RNA in consolidated CHERY plates to prepared 384-well PCR microtitre plate using Hamilton MicroLab STARlet Co-RE 96-channel liquid handler. Combinations of one, two or four targets can be screened per 384-well plate, according to the checkerboard pattern, for which different Hamilton methods are used.
5. Conduct PCR reaction on prepared 384-well PCR microtitre plate using conditions as specified by manufacturer’s protocol.
6. Upload qPCR output to cloud-based platform for analysis.
7. Data quality control and assay performance monitoring.
8. Prepare report for approval.
9. Release report.
10. Epidemiological significance and operational performance data analytics.

#### 2.1.3 LIMS Design

A summary of the in-house developed LIMS solution (ReflX) is as follows:

ReflX is a small, highly customised application written in Python developed to fulfil the requirement for tracking samples end-to-end and provide appropriate levels of traceability. The database is stored in a Microsoft Azure Blob Storage Container, in a comma-separated values format (.csv). Required data input consists of plate maps for each cherry-picked (CHERY) plate being analysed.

Operational data are captured through a series of forms built using PyQt5 (PyQT, 2012) for graphical user interface elements, with customised logic and regular expressions limiting what may be entered into each field. Data recorded in these forms relate to: personnel ID, mastermix batch info, mastermix quality control info, mastermix dispense patterns in each qPCR plate, and sample dispense patterns in each qPCR plate.

ReflX managed the release of raw, unanalysed qPCR data files from the qPCR instruments to the cloud-based data analysis platform, Ugentec FastFinder4.8 *via* GoAnywhere, the managed file transfer (MFT) provider. After analysis, results were released from FastFinder 4.8 and imported into ReflX where plate map data, mastermix dispense patterns, control IDs and sample dispense patterns were used to index the results files and re-format reports to meet specifications.

Finally, ReflX manages integration with existing sequencing pipelines by re-formatting CHERY plate maps and recording chain-of-custody to ensure both compatibility with external partner systems, end-to-end traceability and data integrity.

### 2.2 PCR Reaction Conditions

PCR kits were supplied by ThermoFisher Scientific: The TaqMan SARS-CoV-2 Mutation Research Panel was prepared according to the manufacturer’s instructions for 384-well 10 μL reactions. The design of the TaqMan assay had a FAM-conjugated probe for the mutant allele and a VIC-conjugated probe for the wild-type allele. PCR reactions were performed on Roche LightCycler 480 analysers with reaction conditions set according to manufacturer’s instructions: 60 °C 30 seconds, 50 °C 10 minutes, 95 °C 2 minutes, 45 x [95 °C 3 seconds, 60 °C 30 seconds], 60 °C 30 seconds. FAM and VIC fluorescence for each reaction was read after every amplification cycle.

The following SNP variant mutation assays were evaluated in this study:

- S.Q493R.CAA.CGA, ID no. CVH49P2
- S.K417N.AAG.AAT, ID no. CVFVKJZ
- S.K417T.AAG.ACG, ID no. CVGZE4X
- S.E484K.GAA.AAA, ID no. CVEPRY3
- S.P681R.CCT.CGT, ID no. CVEPRY4

### 2.3 PCR Analysis

Analysis of RT-qPCR data were conducted using FastFinder4.8 (www.ugentec.com/fastfinder), which uses a proprietary AI-driven curve detection and Ct-calling algorithm. A customised genotyping workflow called genotype based on the Ct value of the amplification curves (*i*.*e*. mutant (FAM) or wild-type (VIC) detection was determined as the amplification curve with the lowest Ct value).

### 2.4 Variant Calling

To initially determine diagnostic sensitivity during the evaluation, the Overall Variant Call by the workflow was based on a simple algorithm that assigned variant based on the pattern of wild-type and mutant allele detection by the individual TaqMan SNP assays. The algorithm for the four-target assay configuration (P681R, E484K, K417N and K417T) is shown in table 1; while the single-target assay configuration algorithm is shown in table 2.

**Table 1:** Calling algorithm for the four-target assay configuration.

| SNP Assay Calling |  |  |  | Overall Variant Call |
| --- | --- | --- | --- | --- |
| P681R | E484K | K417N | K417T |  |
| wild-type | wild-type | wild-type | wild-type | wild-type |
| wild-type | mutant | mutant | wild-type | B.1.351, Beta Variant |
| wild-type | mutant | wild-type | mutant | P.1, Gamma Variant |
| mutant | wild-type | wild-type | wild-type | B.1.617.2, Delta Variant |

**Table 2:** Calling algorithm for the single target assay configuration.

| Assay | Overall Variant Call |
| --- | --- |
| Q493R |  |
| mutant | B.1.1.529/BA.1, Omicron Variant |
| wild-type | NOT B.1.1.529/BA.1, Omicron Variant |

### 2.5 Control Material

For validation and routine use, control materials were supplied by Twist Bioscience. The following synthetic RNA controls were used:

- Control 16, B.1.351, Beta Variant
- Control 17, P.1, Gamma Variant
- Control 23, B.1.617.2, Delta Variant
- Control 48, B.1.1.529/BA.1, Omicron Variant

In routine use, this material was diluted and aliquoted from 10^9^ copies mL^*-*1^ stock to 2 × 10^3^ copies mL^*-*1^ working concentration.

In addition, the following inactivated virus controls were used for validation only:

- SARS-CoV-2 WHO First International Standard (NIBSC)
- SARS-CoV-2 B.1.351, Beta Variant (NIBSC)
- SARS-CoV-2 B.1.617.2, Delta Variant (NIBSC)
- SARS-CoV-2 P.1, Gamma Variant (NIBSC)
- RTX1 Q Control Panel (Qnostics)
- RTX2 Q Control Panel (Qnostics)
- RTX3 Q Control Panel (Qnostics)
- RTX4 Q Control Panel (Qnostics)
- RTX5 Q Control Panel (Qnostics)
- RTX Medium Q Control Panel (Qnostics)
- AccuPlex SARS-CoV-2 Verification Panel - Full Genome (SeraCare)

### 2.6 Data Analysis

Validation and research data were analysed in R (R Core Team, 2022) and figures produced using the package ggplot2 (Wickham, 2016).

### 2.7 Sample Collection

Patient samples were obtained from the Department of Health and Social Care (DHSC) Test and Trace programme, one purpose of which was to support public health surveillance and wider clinical decision-making. Samples were anonymised on collection by an eleven character barcode, which was recorded in the national system. The anonymised barcode was subsequently recorded in the ReflX database, and as such the data collected in the ReflX database could not be traced back to individuals.

## 3 Results

### 3.1 The ReflX workflow is both highly sensitive and specific for the detection of Variants of Concern

Initially, four TaqMan assays (P681R, E484K, K417N, K417T) were assessed against SARS-CoV-2 wild-type (NIBSC reference strain First WHO International Standard for SARS-CoV-2) and SARS-CoV-2 variant (NIBSC SARS-CoV-2 Beta, Gamma and Delta) material. The Q493R assay was not included in initial experimentation and was only introduced late in the evaluation in response to the rise in B.1.1.529/BA.1 (Omicron) cases. For the Beta variant, the expected SNPs were E484K and K417T. For the Delta variant, only the P681R mutation should be detected. For the Gamma variant, it is possible for both K417T and K417N to be detected. Where the mutation is not detected, the result should be returned as wild-type (*i*.*e*. P681P, E484E, K417K). The results, presented in table 3 show that all four SNP TaqMan assays correctly called the wild-type or mutant alleles, and hence showed specificity for the tested variants.

**Table 3:**
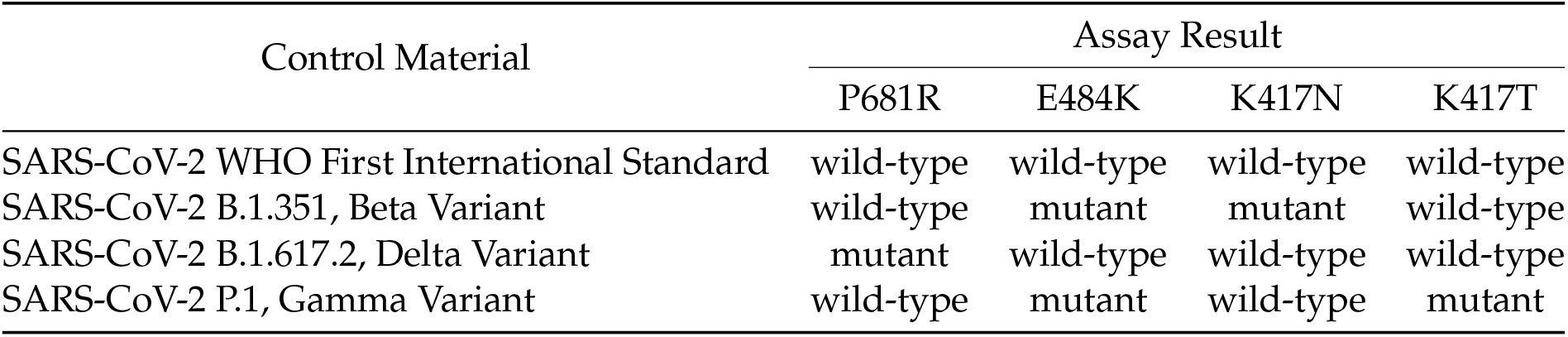
Specificity analysis of the P681R, E484K, K417N and K417T mutation assays against inactivated wild-type and variant control materials supplied by NIBSC.

| Control Material | Assay Result |  |  |  |
| --- | --- | --- | --- | --- |
|  | P681R | E484K | K417N | K417T |
| SARS-CoV-2 WHO First International Standard | wild-type | wild-type | wild-type | wild-type |
| SARS-CoV-2 B.1.351, Beta Variant | wild-type | mutant | mutant | wild-type |
| SARS-CoV-2 B.1.617.2, Delta Variant | mutant | wild-type | wild-type | wild-type |
| SARS-CoV-2 P.1, Gamma Variant | wild-type | mutant | wild-type | mutant |

Next, to evaluate specificity of the assays against common respiratory pathogens, the assays were screened against five Qnostics Q Control Respiratory Pathogen Panels (Qnostics RTX1 Q Control, Qnostics RTX2 Q Control, Qnostics RTX3 Q Control, Qnostics RTX4 Q Control, Qnostics RTX5 Q Control and RTX Medium Q Control). No false wild-type or mutation results were produced from testing these panels, indicating that the assays are not cross-reactive with the twenty tested respiratory pathogens (table 4).

**Table 4:** Specificity analysis of the P681R, E484K, K417N and K417T mutation assays against a panel of respiratory pathogens. No amplification = na.

| Respiratory Pathogen Target | Assay Result |  |  |  |
| --- | --- | --- | --- | --- |
|  | P681R | E484K | K417N | K417T |
| SARS-CoV-2 | wild-type | wild-type | wild-type | wild-type |
| Influenza A H1N1 | na | na | na | na |
| Influenza B Victoria | na | na | na | na |
| Respiratory Syncytial Virus Type A | na | na | na | na |
| Respiratory Syncytial Virus Type B | na | na | na | na |
| Adenovirus Type 1 | na | na | na | na |
| Adenovirus Type 14 | na | na | na | na |
| Rhinovirus Type 16 | na | na | na | na |
| Metapneumovirus Type A2 | na | na | na | na |
| Parainfluenza Virus Type 1 | na | na | na | na |
| Parainfluenza Virus Type 2 | na | na | na | na |
| Parainfluenza Virus Type 3 | na | na | na | na |
| Parainfluenza Virus Type 4 | na | na | na | na |
| Coronavirus Type 229E | na | na | na | na |
| Coronavirus Type NL63 | na | na | na | na |
| Coronavirus Type OC43 | na | na | na | na |
| Mycoplasma pneumoniae | na | na | na | na |
| Enterovirus Type A16 | na | na | na | na |
| Legionella pneumophila | na | na | na | na |
| Enterovirus Type 68 | na | na | na | na |

In addition, to ensure that the detection of correct allele occurs in a mixed matrix, SARS-CoV-2 wild-type and variant material were combined with a human buccal swab and an RTX Q Control Panel. The results of this experiment confirm that the assays have: a) no cross-reactivity with either human nucleic acid or with a panel of respiratory pathogens, and b) can specifically detect the correct allele in a simulated patient sample carrying a co-infection (table 5).

**Table 5:**
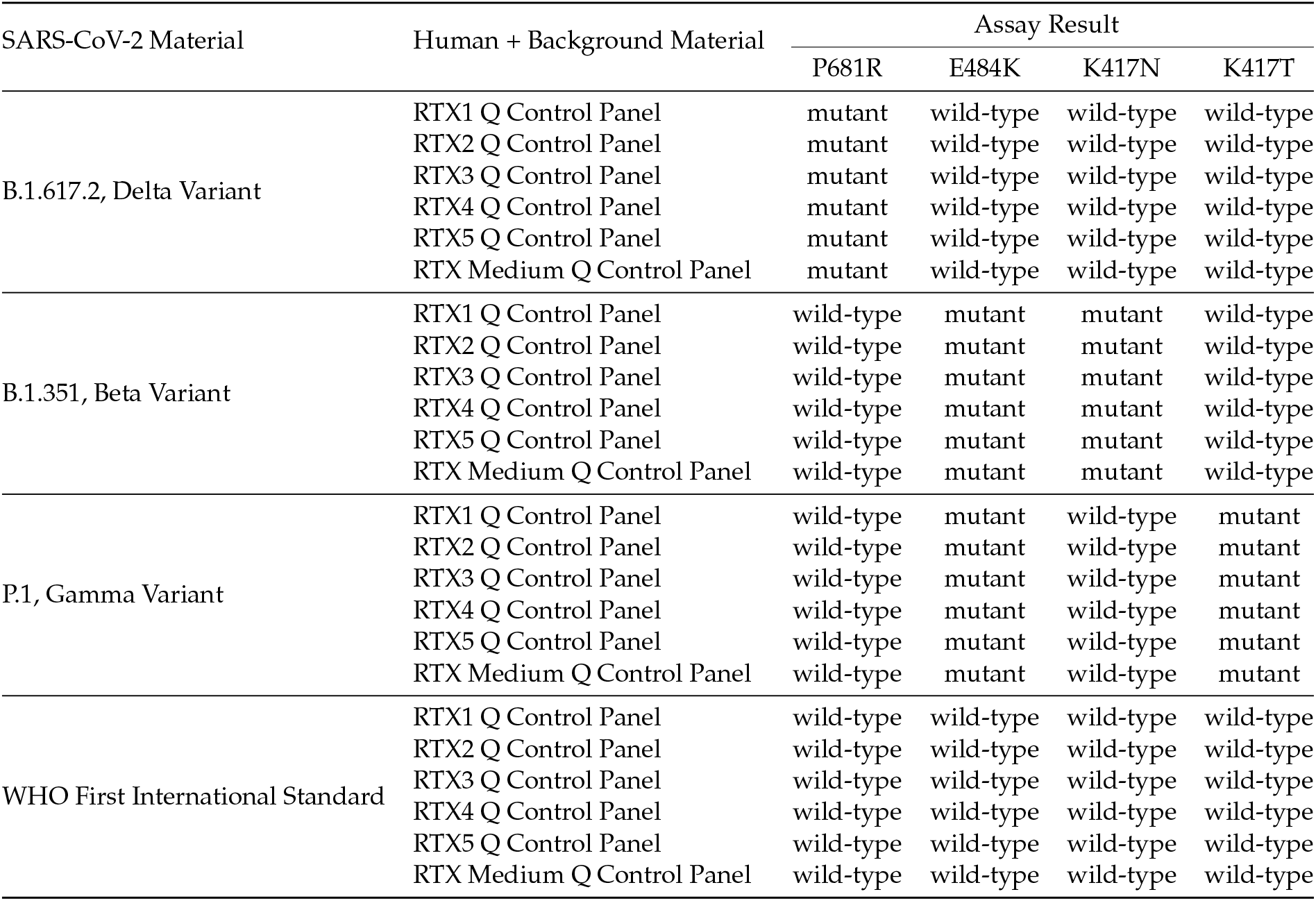
Confirmation of specificity in background of other respiratory pathogens and human material. SARS-CoV-2 material was mixed with RTX Q Control Panels along with a human buccal swab to simulate a real co-infected patient sample.

For evaluation of the analytical sensitivity of the assay for the mutant alleles, a 2-fold dilution series of Twist Bioscience synthetic RNA SARS-CoV-2 variant reference material was prepared starting from the pre-extraction equivalent of 15,000 copies mL^*-*1^. Eight concentrations of each of the variant reference materials were tested 24 times against the genotyping assays to determine the detection rate. The lower limit of detection was determined by logit analysis and is, by convention, the concentration at which there is a 95% probability of detection of the analyte (LLoD95). Note that during the evaluation, the Omicron variant, B.1.1.529/BA.1 emerged and increased dramatically in prevalence hence the inclusion of the Q493R SNP TaqMan assay in the subsequent experimentation. The calculated LLoD95 for the mutant alleles are summarised in table 6. The LLoD95 for the wild-type alleles was estimated from a dilution series prepared from AccuPlex SARS-CoV-2 Verification Panel - Full Genome. In this experiment, n = 4 due to limitations in quantity of starting material, hence a logit analysis could not be reliably conducted and a range in which the LLoD95 is likely to fall is summarised in table 7. These data show that the limits of detection for the mutant alleles fell comfortably below 1000 copies mL^*-*1^, and while the limit of detection could not be accurately determined for the wild-type allele with this experiment, it is likely to fall below 2500 copies mL^*-*1^.

**Table 6:** Analytical sensitivity evaluation for mutant allele targets. Lower limit of detection (LLoD95) was determined through logit analysis of a dilution series, n = 24.

| TaqMan Assay | LLoD95 (copies mL<sup>-1</sup>) |
| --- | --- |
| P681R | 275 |
| E484K | 457 |
| K417N | 441 |
| K417T | 545 |
| Q493R | 501 |

**Table 7:** Analytical sensitivity range estimates for wild-type allele targets, estimated from a dilution series, n = 4.

| TaqMan Assay | Limit of Detection Range (copies mL<sup>-1</sup>) |
| --- | --- |
| P681P | 1250 - 2500 |
| E484E | 313 - 625 |
| K417K | 1250 - 2500 |
| K417K | 625 - 1250 |
| Q493R | 1250 - 2500 |

### 3.2 The ReflX workflow is highly concordant with Whole Genome Sequencing

For the evaluation of diagnostic sensitivity and specificity, 159 patient samples that had been called as detected for SARS-CoV-2 in the LGC Biosearch Technologies End-Point PCR system were screened against the variant targets. 77 of these patient samples were screened against the P681R, E484K, K417N and K417T assays, while the set of 82 remaining patient samples were screened against the Q493R assay. All samples were subsequently processed through a WGS pipeline to confirm the lineage, with the WGS result assumed to be final. Concordance of the genotyping assay and WGS was calculated using overall percentage agreement (OPA), defined as the frequency of the variant being correctly called according to the mutant and wild-type allele detection by the genotyping assay, and confirmed by WGS. Samples that did not return a WGS result, or had neither a genotyping or sequencing result were excluded from the analysis (33 of the 77 samples in the P681R, E484K, K417N and K417T cohort, and 5 of the 82 samples in the Q493R cohort). The results from this experiment are summarised in table 8. There was a single ‘Not Concordant’ result associated with the Q493R assay, which was for a sample where there was a WGS result available, but an ‘Invalid’ Genotyping result due to no amplification. Such a result may have occurred due to low volume in the source plate, or low concentration causing stochasticity in result detection. Overall, the outcome of the concordance study suggest that in terms of variant calling, the ReflX genotyping workflow gives a comparable end result to WGS, with an overall percentage agreement of 100% for the four-target assay, and 98.7% for the single-target assay.

**Table 8:** Concordance between Genotyping call and WGS. Samples were excluded from the Overall Percentage Agreement if there was no WGS data available to concord with.

|  | Assay Configuration |  |
| --- | --- | --- |
|  | P681R, E484K, K417N, K417T | Q493R |
| No WGS Data | 30 | 3 |
| No Genotyping or WGS Data | 13 | 2 |
| Not Concordant | 0 | 1 |
| Concordant | 34 | 76 |
| Overall Percentage Agreement | 100% | 98.7% |

### 3.3 ReflX tracked the emergence of Omicron Sub-lineages BA.4 and BA.5 in the UK

In late November 2021, the UKHSA declared the original Omicron lineage, B.1.1.529/BA.1 as a variant under investigation (VUI). In late January 2022, B.1.1.529/BA.1 had become the dominant strain in the UK, outcompeting the B.1.617.2 (Delta) variant. At the same time, cases of BA.2, a sub-lineage of Omicron, were beginning to increase. Following this, several Omicron recombinants, including XD, XE and XF, were detected by the WGS program, but as they belonged to the Omicron lineage they all retained the Q493R mutant allele. In early May 2022, cases of Omicron BA.4 and BA.5 started to be detected in WGS. BA.4 and BA.5 carry a reversion of the mutation at the Q493 locus to the wild-type, Q493Q. The emergence and rise in BA.4 and BA.5 cases coincided with an observed increase in wild-type allele results in the single-target Q493R ReflX workflow (figure 1). The earliest indication of the emergence of the BA.4 and BA.5 sub-lineages can be traced to mid April where a small but perceptible rise in wild-type calls was detected. This trend continued over the course of approximately two months and by July the vast majority of SARS-CoV-2 detected patient samples carried the wild-type Q493Q form of the allele.

**Figure 1:**
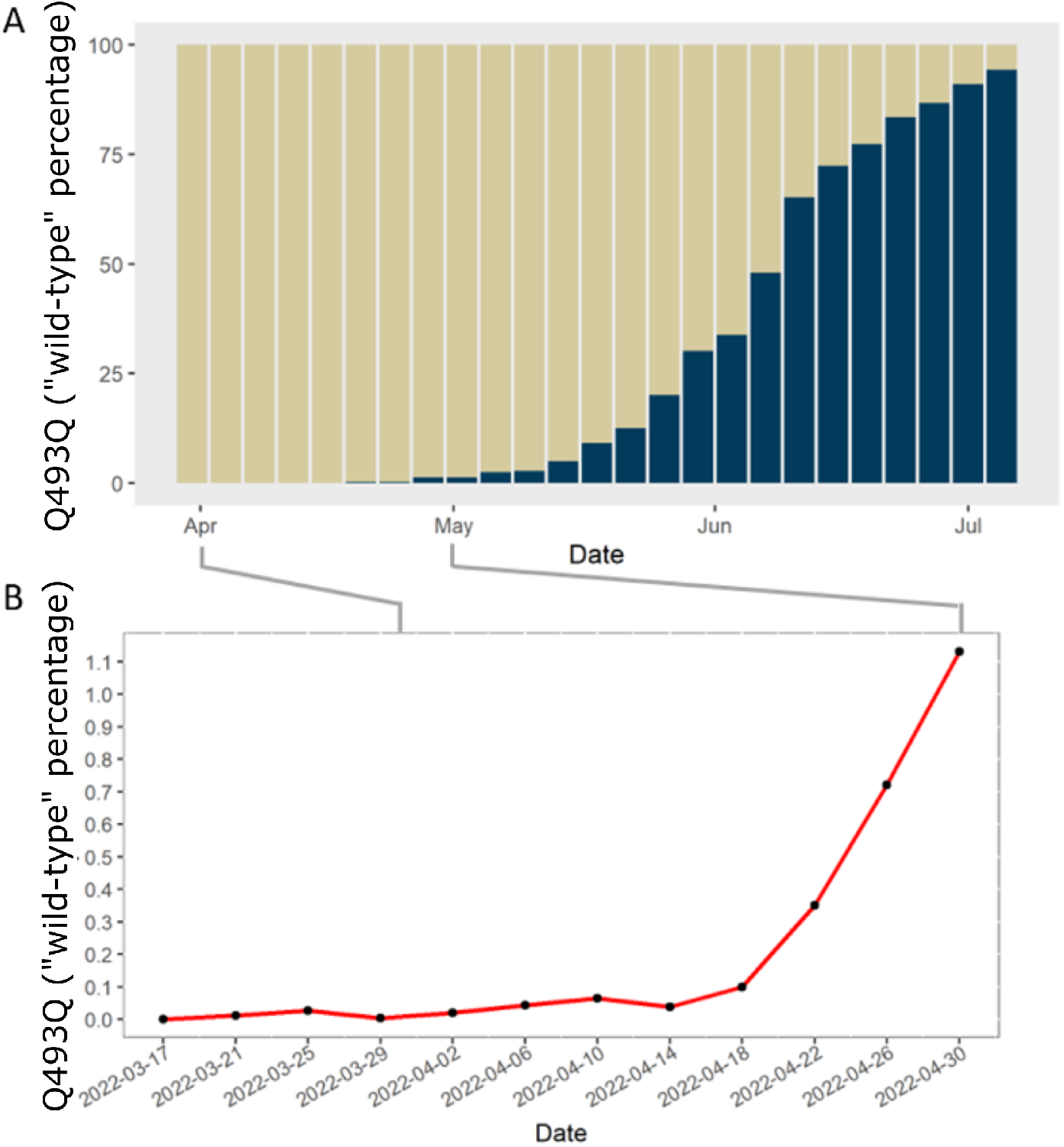
Between April and July 2022 the ReflX system tracked a concurrent reduction in the Q493R mutant allele and increase in the Q493Q wild-type allele to give a real-time indication of the prevalence of BA.4 and BA.5 in samples taken from the testing program in the United Kingdom. Note: the x-axes are the date that the result was released by the laboratory, (typically 24 - 48 hours after the swab was taken). A) Proportional stacked bar plot of Q493 locus. Sand = proportion of Q493R mutant allele, Navy = Q493Q wild-type allele. B) Expanded axis shows that as early as 22^*nd*^ April an increase in the prevalence of the Q493Q wild-type allele was detected by the ReflX system.

## 4 Discussion

Emerging SARS-CoV-2 variants have prolonged the duration of the COVID-19 pandemic due to their potential for vaccine escape and increased transmissibility (Tao et al., 2021). It is widely acknowledged that rapid high throughput diagnostics are required to effectively track and manage a rapidly mutating pathogen at a population scale. More sophisticated monitoring assesses how the infectious agent is evolving, which in turn informs public health strategies (Knyazev et al., 2022). WGS is the gold standard for characterising novel SARS-CoV-2 lineages, but is expensive and time-consuming, creating a barrier to entry for laboratories and health-care services without existing infrastructure. PCR-based genotyping cannot be used to detect novel mutations, but can be used to screen for known mutations and is comparatively quick, inexpensive and can be carried out using basic molecular biology equipment. An additional advantage of PCR-based genotyping is that both the lab work and the analysis can be easily automated for processing high sample volumes. Whilst genotyping approaches have been tried at relatively low sample volume by multiple groups previously (Mertens et al., 2022; Nörz et al., 2021a; Nörz et al., 2021b), here we report the operationalisation of a scalable partially automated system designed to screen up to 36,000 samples in a 24-hour period, based on eight qPCR instruments and a 10% positivity rate. From its launch at the end of December 2021 until its retirement in late 2022 (due to the ramp-down of SARS-CoV-2 testing aligned with the UK government’s ‘Living with COVID’ strategy (Cabinet Office, 2022)), the ReflX service was run daily and was responsible for genotyping *c*. 400,000 SARS-CoV-2 samples, thereby demonstrating its robustness and scalability.

Five commercially available TaqMan mutation assays were assessed for routine use, which were P681R, K417N, K417T, E484K and Q493R. The evaluated assay SNP targets are associated with the Beta, Delta, Gamma, and Omicron SARS-CoV-2 variants and were highly sensitive and specific even in the presence of common respiratory pathogens. Additionally, all five targets were confirmed to have a high degree of concordance with WGS, with the four target (P681R, K417N, K417T, E484K) assay configuration achieving 100% concordance, and the Q493R single target assay configuration 99% concordance. However, at the time of assay evaluation, Beta and Gamma variants were no longer in wide circulation, thus the mutant probes for targets E484K and K417T could not be interrogated during the diagnostic sensitivity and specificity evaluations.

PCR assays have been useful in analysing the emergence and spread of new SARS-CoV-2 lineages in the United Kingdom on at least three separate occasions. Firstly, the widely-reported ‘S-Gene Target Failure’ (SGTF) was a consequence of the spike .6 H69/V70 deletion in the Alpha variant that resulted in increased infectivity (Meng et al., 2021) and viral loads (Kidd et al., 2021). SGTF was also a proxy for the spread of the Omicron variant (BA.1) as it replaced the Delta variant (Paton et al., 2022), while our data presented here in figure 1 show the replacement of BA.2 by lineages carrying the Q493Q wild-type allele, later confirmed by WGS to be BA.4 and BA.5. It is important to note that genotyping results are only indicative of the lineage to which a sample belongs, a definitive variant call would require WGS, hence determination of which new variant is driving a trend would need to await WGS confirmation. An organisation planning an infectious agent surveillance programme would therefore do well to maintain communication channels and data-pipelines between labs performing sequencing and labs performing genotyping as their activities are complementary.

The spike protein is under constant selective pressure due to its importance in pathogenicity and being a target for diagnostics and therapy. The Omicron variant carries conformation changes in the Receptor Binding Domain (RBD) of the spike protein, which are hypothesised to restrict access to previously exposed epitopes (Zhao et al., 2022). It may be possible to design a SNP panel that screens mutations around and within the RBD, as the intensity of selection pressure at this location means it holds a high potential for mutations that evade the immune system. Interestingly, there have also been some mutant alleles that have arisen in variants and then reverted to their ancestral wild-type allele in sub-lineages, whether through recombination or mutation giving a selective advantage *e*.*g*. .6 H69/V70, Q493R, K417N, N440K, G142D, N764K (Qu et al., 2022; Philip et al., 2023; Zhao et al., 2023). A horizon-scanning application for a genotyping assay therefore could leverage genomic regions of concern, such as RBD; alongside such reversible SNPs to horizon-scan for the emergence of mutations.

In the absence of multiplexing, the design of the ReflX workflow (or any PCR-based genotyping workflow) would mean that should the number of SNP targets increase, throughput would decrease (*e*.*g*. 4 wells would be occupied by 4 samples tested against one target, versus 4 wells occupied by 1 sample tested against 4 targets). Therefore, if an organisation were to use genotyping for horizon-scanning surveillance for multiple mutations, pooled samples should be considered as a strategy to increase throughput (and reduce cost). Pooling and cherry-picking strategies should also consider sample meta data such as geography in the workflow algorithms to help identify epidemiological patterns.

A single target genotyping assay is admittedly of limited use when it comes to monitoring for the emergence of variants, so it was serendipitous that the BA.4 and BA.5 sub-lineages of Omicron carried the Q493Q wild-type allele, which allowed for their differentiation from BA.1 and BA.2 by the Q493R assay. Nevertheless, the ReflX workflow was developed to be adaptable to the needs of the moment, and at the core of the design is the balance between a completely locked-down system that is rigid and unchangeable, and an undefined free system that is completely flexible, but also uncontrolled. To achieve this, boundaries to the flexibility had to be set: The overall workflow pattern and order was inflexible with the steps consisting of Cherry-Picking, Mastermix and PCR plate preparation, template addition, PCR reaction, and finally analysis. However, within the steps there were interchangeable components, including: PCR mastermix reagents, positive controls and the checkerboard patterns on the PCR plates. This design philosophy is similar to the principles underpinning an ISO15189 flexible scope accreditation: whereby a pathology laboratory will set the boundaries of its flexible scope, and demonstrate its technical competence within said boundaries to a regulator. The laboratory is then in turn permitted to validate and deploy new assays without requiring an extension to scope application to the regulatory body (Thelen et al., 2015; UKAS, 2019).

The most efficient laboratory workflows are highly customised and tailored to the application. Indeed, off-the-shelf solutions are rarely perfect immediately and require at least some configuration to meet end user requirements and maximise productivity. Yet, it is not hard to foresee the advantages that a flexible LIMS and workflow solution (within certain boundaries) could have in preparation for the emergence of a new pandemic-causing infectious agent. Assuming that the infectious agent is microbial or viral in nature, a PCR-based assay is likely to be utilised in the first instance as a primary diagnostic tool. Hence, the overall system for testing will be largely identical to existing PCR workflows, just with interchangeable components, which would be fortuitous should there be supply chain challenges in the early stages of ramping up of production of diagnostic reagents and consumables.

In conclusion, the ReflX workflow presented here has the capability to support a flexible SARS-CoV-2 genotyping service and is a complete sample-to-result solution for high volume patient testing. Streamlined sensitivity/specificity studies and a sensible quality management system coupled with the flexible design of the LIMS and robotics logic mean that the workflow can be rapidly adapted to screen for alternative targets from novel SARS-CoV-2 variants. While the threat to human health by COVID-19 has been largely mitigated through the successful roll-out of mass-vaccination campaigns, the risk of the emergence of a novel vaccine escape variant remains. Now that the focus turns to developing pandemic preparedness strategies for ‘Pathogen X’ (Skyle, 2022), it would be reasonable to also create strategic pandemic-ready diagnostic workflow solutions that strike the delicate balance between control and flexibility.

## Data Availability

All data produced in the present study are available upon approval of a freedom of information request to the UK Health Security Agency (UKHSA).

## 5 Notes

This work was financially supported by the UK Department of Health and Social Care through the Test and Trace programme. The work was conducted at the Rosalind Franklin Laboratory (Royal Leamington Spa, UK). As all data were obtained from anonymised patient samples routinely collected as part of the Test and Trace programme, there were no prospective samples collected. The use of samples for genotyping as described in this work is covered by the over-arching Privacy Notice issued by the UKHSA with the legal basis for processing such data being:

- GDPR Article 6(1)(e) – the processing is necessary for the performance of its official tasks carried out in the public interest in providing and managing a health service
- GDPR Article 9(2)(h) – the processing is necessary for the management of health/social care systems or services
- GDPR Article 9(2)(i) – the processing is necessary for reasons of public interest in the area of public health
- Data Protection Act 2018 – Schedule 1, Part 1, (2) (2) (f) – health or social care purposes

